# A Measurement-Based Care Strategy for Buprenorphine-Naloxone Treatment (Bup-MBC): Development of an EHR-Integrated Intervention

**DOI:** 10.64898/2026.08.27.26361539

**Authors:** Thomas J. Reese, Carolyn M. Audet, Jessica S. Ancker, Adam Wright, David E. Marcovitz, Kristopher A. Kast, John F. P. Bridges, Hilary A. Tindle, Mauli V. Shah, Amanda von Horn, Michael Matheny

## Abstract

**Introduction:** Risk of recurrent opioid use during buprenorphine-naloxone (bup-nx) treatment is dynamic and remains elevated after initiation, with vulnerability shaped in part by treatment intensity and gaps between visits, yet routine outpatient care relies on episodic encounters and retrospective data. This mismatch can delay recognition of emerging instability and limit timely treatment adjustments. This paper reports the development and specification of an intervention strategy to address this mismatch.

**Methods:** We used a structured, multi-phase design process to specify and configure a measurement-based care (MBC) strategy for bup-nx treatment (Bup-MBC) in outpatient addiction clinics through three phases: (1) a systematic review of patient-reported outcome measures (PROMs) for substance use treatment; (2) a qualitative needs assessment using the Theoretical Domains Framework and COM-B (Capability, Opportunity, Motivation–Behavior) model to identify gaps in risk monitoring, agency, and trust; and (3) iterative co-design with multidisciplinary clinicians to refine workflow fit and trust-preserving use of data. Patients informed item and feedback content during the needs assessment but did not participate in the co-design cycles.

**Results:** Bup-MBC integrates (1) brief between-visit PROMs (e.g., withdrawal, craving, adherence); (2) immediate non-punitive patient feedback; (3) clinician-facing summaries and non-directive prompts in the electronic health record (EHR); and (4) an opt-in between-visit outreach pathway with predefined safety triggers, all configured within existing EHR and patient portal infrastructure. It targets patient and clinician capability to recognize changes in risk, opportunity for action through structured monitoring and visit preparation, and trust and agency through non-punitive communication, without adding substantial burden. The full measure set, severity bands, and question-to-action map are provided as supplementary material. Key trade-offs included prioritizing single-item measures for feasibility, balancing opt-in outreach with safety overrides, and assuming routine clinician use of summaries.

**Conclusion:** This development study specifies an EHR-integrated MBC strategy for outpatient bup-nx treatment. As single-center design work with co-design limited to clinicians and delivery contingent on portal or text-message access, its outputs are hypotheses about mechanism and fit rather than demonstrated effects. Feasibility studies are needed to evaluate uptake, acceptability, workflow fit, and effects on treatment.

## INTRODUCTION

Risk of recurrent opioid use during buprenorphine naloxone (bup nx) treatment is dynamic, with many patients resuming illicit or non-prescribed use within months of initiation and vulnerability often shifting between visits and during transitions in treatment intensity, such as changes in visit frequency or level of care.^1–3^ Routine outpatient bup nx care is typically organized around episodic encounters and retrospective indicators such as urine drug screening and crisis driven contact, which provide delayed, intermittent snapshots of use and may not capture emerging instability between visits.^4–6^

A prior qualitative study of patients receiving bup nx and their providers found that the mismatch between continuous recurrent risk and intermittent monitoring created tensions between preventing recurrent use and supporting patients’ treatment agency.^7^ In particular, these tensions arose in how monitoring was framed and how decisions were made during and between visits. Patients described informal self-monitoring of withdrawal, craving, and triggers without structured feedback or shared interpretation, while clinicians reported relying on intuition, using only sparse data points to guide dosing and visit frequency. Monitoring practices were experienced as either supportive or punitive depending on their timing, transparency, and how information was used in subsequent decisions about medication and visits. Organizing these findings with the COM B (Capability, Opportunity, Motivation – Behavior) framework highlighted three challenges in existing care: limited shared understanding of evolving risk (capability), few timely, systematic signals between visits (opportunity), and fragile trust and agency in the face of potential punishment (motivation). Together, these gaps underscored the need for systematic, trust preserving ways to detect changes in risk between visits and guide responsive adjustments in care.^8^ COM B was used as a practical framework for linking empirically identified barriers to modifiable targets for intervention design.

Measurement-based care (MBC) uses brief patient-reported outcome measures (PROMs) to track symptoms and functioning over time and guide clinical decisions, and it has improved engagement and outcomes in multiple behavioral health settings.^9–11^ Emerging work is beginning to apply MBC to opioid use disorder (OUD), including approaches that use DSM-5 symptoms to monitor response to medications, but these efforts largely emphasize symptom counts and retention rather than the relational and agency-sensitive aspects of bup-nx treatment.^12,13^ Moreover, direct evidence that MBC improves treatment outcomes in OUD remains limited, underscoring the need for transparent development and staged evaluation of MBC strategies in this population.^13,14^ In principle, MBC could address the gaps from our needs assessment by making between-visit fluctuations visible, creating shared information, and structuring balanced responses to emerging recurrent use risk.^7,9^ In OUD care, where stigma and fears of punitive monitoring are common, generic MBC models may feel like surveillance or algorithmic control rather than support unless they are deliberately designed to preserve trust and agency.^15^

In this work, bup-nx was the evidence based treatment, and MBC served as an implementation strategy to support safer and more patient centered use.^16,17^ We used a theory guided needs assessment with iterative co design, to specify and integrate a pragmatic, technology enabled MBC strategy that addresses specific behavioral and relational gaps in outpatient bup nx care. Building on a systematic review of single item patient reported outcome measures, a qualitative needs assessment, and iterative clinician co design, the strategy integrates brief between visit patient reported measures, automated patient facing feedback, clinician facing EHR summaries, and an opt in pathway for between visit outreach, all configured within the EHR and patient portal/text messaging infrastructure to preserve trust and agency while enabling timely responses to recurrent risk.^18–23^ The objective of this paper is to transparently report the development and specification of an implementation strategy to enhance the uptake of bup-nx treatment. Specifically, we aim to (1) describe the design process that shaped the strategy, (2) specify the strategy content, and (3) articulate the hypothesized mechanisms, assumptions, and boundary conditions to be tested in subsequent studies.

## METHODS

### Design and theoretical framework

We used a co-design process to design and configure an MBC implementation strategy for bup-nx treatment (Bup-MBC). The process proceeded in three phases that informed the final intervention specification now embedded in the EHR and patient portal. First, we conducted a systematic review of single-item PROMs for substance use treatment to identify constructs and instruments suitable for routine MBC monitoring.

Second, we carried out a qualitative needs assessment using the Theoretical Domains Framework (TDF) and the COM-B model to characterize gaps in recurrent use risk monitoring, agency, and trust in bup-nx treatment. Third, we held iterative co-design sessions and pilot testing with multidisciplinary clinicians in the target clinics to refine draft Bup-MBC components with respect to item wording, burden, workflow integration, and trust-preserving use of data.^7,18–24^ We use “trust-preserving” to describe monitoring practices that patients experience as supportive rather than punitive, as characterized in our needs assessment by transparency about how data are used, non-judgmental framing, and decisions made with the patient; this working definition was derived from patient and clinician accounts.^7^ In the needs assessment, empirically derived determinants such as withdrawal avoidance, trust-sensitive monitoring, and stigma-driven ambivalence were mapped onto COM-B components, and Bup-MBC was designed to target these determinants using aligned behavior change techniques. In selecting constructs, items, and workflows, we drew on prior PROM and implementation work in substance use and recovery oriented care, including studies on PROM use in substance use disorder services and recovery focused outcomes.^12,25–28^ Our reporting follows the Template for Intervention Description and Replication (TIDieR) to ensure sufficient detail regarding rationale, materials, procedures, and delivery, and **Table 2** provides a TIDieR-aligned summary of Bup-MBC as currently configured for pilot implementation.^29^ Because patient involvement is central to claims of human-centered design, we additionally report the nature and limits of patient input, consistent with reporting guidance.^30^ The complete measure set and question-to-action map are provided in **Supplement Tables 1 and 2** to support reproduction and adaptation.

### Ethics approval

The qualitative needs assessment (patient and clinician interviews) and the clinician co-design activities were reviewed and approved by the Vanderbilt University Medical Center Institutional Review Board (IRB #231316). Configuration and technical testing of Bup-MBC in the EHR used test-patient records only and involved no identifiable patient data.

### Formative co-design and pilot refinement

We conducted iterative co design and pilot refinement with clinicians practicing in outpatient addiction psychiatry clinics at Vanderbilt University Medical Center. Purposive sampling was used to recruit providers, social workers, and recovery coaches who provided bup-nx care in the target clinics (n = 6 clinicians: 3 providers [i.e., 2 attending and 1 fellow in addiction psychiatry], 1 social worker, and 2 recovery coaches), inviting them via email and clinic meetings to participate in brief design sessions. In each of the three cycles over seven months, clinicians reviewed draft items, patient facing messages, and EHR summary mock ups and provided structured feedback focused on item wording and recall period, perceived burden, workflow fit, and trust preserving use of data. We captured suggested changes on a living “question to action” map that linked each response option to (a) patient facing feedback, (b) branching prompts for contextual detail, (c) severity bands, and (d) outreach/workflow routing. After each cycle, the study team revised the map and associated artifacts, which were then re reviewed in subsequent sessions until no new substantive changes were suggested and clinicians agreed that the content and workflows were acceptable and feasible for pilot use.

Sessions lasted approximately 45 minutes and were held by videoconference. Artifacts including the mock-ups and question-to-action map were provided prior to the session for review. Feedback was captured in real time during sessions using a structured template organized by intervention component, and additional written feedback was provided afterwards on the artifacts themselves. After each cycle two coauthors (TJR and MVS) reviewed the logged feedback and proposed dispositions about whether and how to implement changes with study team clinicians (KAK and DEM), prioritizing patient safety first then trust preservation and workflow feasibility. A change was considered substantive if it altered item content, response scaling, severity-band boundaries, feedback language, or workflow routing, as distinct from cosmetic wording or formatting adjustments; iteration stopped when a complete cycle generated no new substantive change requests. Pilot testing in this phase consisted of structured walkthroughs of the configured EHR and patient-portal build using test-patient records, in which clinicians completed PROMs as patients would, viewed the automated feedback, and reviewed the resulting EHR summaries. Patients did not participate directly in these co-design cycles. Patient input into intervention content occurred earlier during the needs-assessment interviews when patients reacted to draft items and feedback messages. This input was carried into the co-design sessions.

### Setting and context

The intervention targets outpatient bup-nx treatment in addiction psychiatry clinics at Vanderbilt University Medical Center, where care is delivered by multidisciplinary teams (providers, social workers, recovery coaches) using established EHR and patient portal/text messaging infrastructure. ^31–33^ The current implementation uses team based care models where providers are supported by other clinicians for outreach and follow up. Bup-MBC will be offered to adults (≥18 years) initiating bup-nx treatment for OUD who are able to receive secure messages and complete assessments in English. During the planned pilot, patients seen by a participating provider will be offered enrollment, and the denominator of eligible patients, the proportion reached, and the characteristics of reached versus not-reached patients will be reported as feasibility outcomes.

### Intervention components and design goals

Bup-MBC consists of four integrated components currently configured within the EHR and patient portal:

(1) structured patient reported outcome monitoring, (2) automated patient feedback and self reflection, (3) clinician facing summaries and decision support, and (4) an opt in between visit outreach pathway. **Figure 1** shows the computer version (versus smart phone) of the patient interface for between-visit PROM completion and immediate feedback. Bup-MBC targets a limited set of patient and clinician behaviors identified as leverage points and is intended to complement, not replace, clinical judgment and therapeutic relationships. (**Box 1**). Design goals are to improve patients’ and clinicians’ shared capability to understand symptoms and risk, opportunity for action through structured between visit monitoring and visit preparation, and trust and patient agency through non punitive communication, reduced fear of punishment, and reinforcement of patients’ participation in decisions, without adding substantial workload or introducing new punitive thresholds.

**Figure 1.**
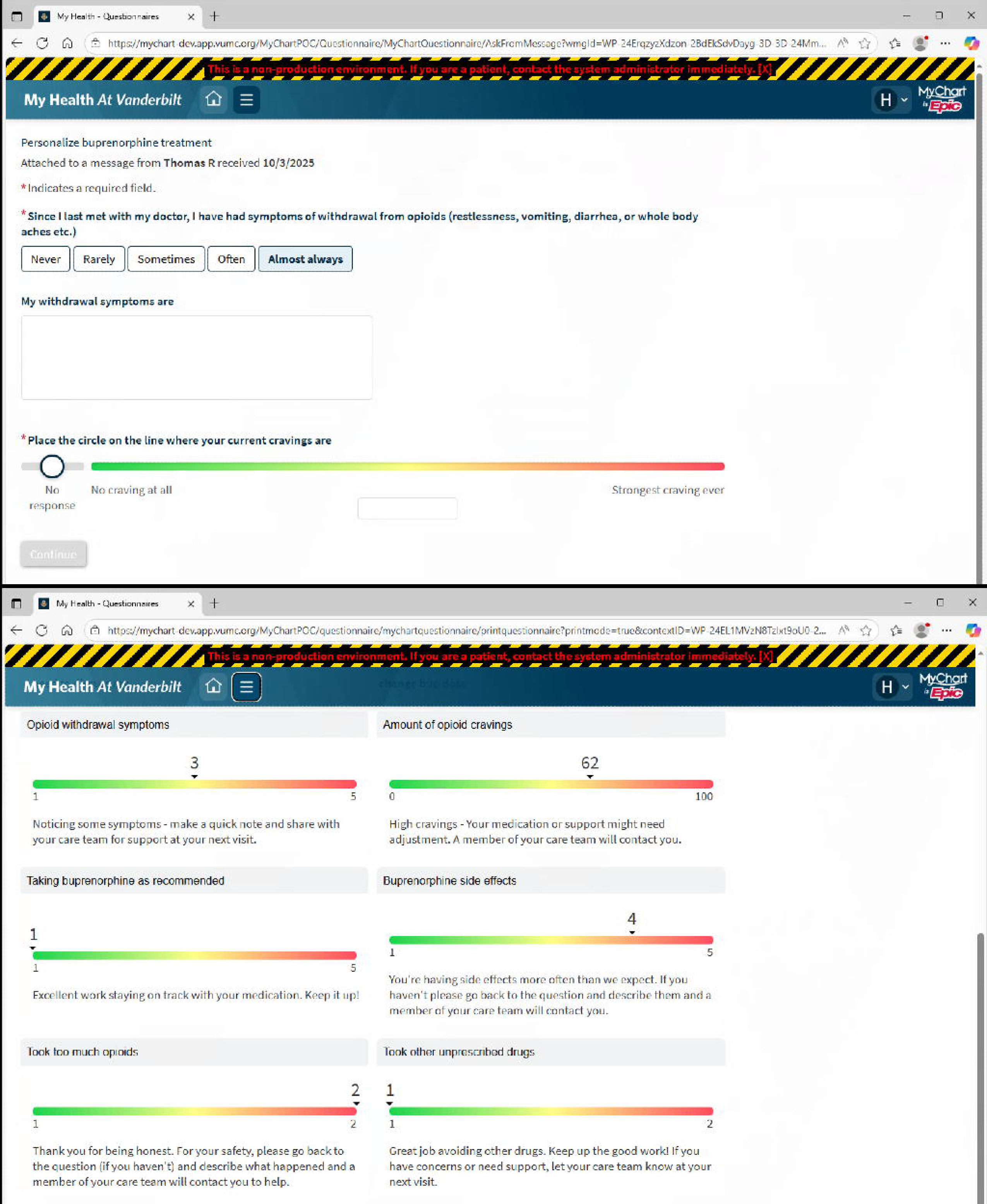
Patient interface for between visit PROM completion and immediate, non punitive feedback.

**Box 1.**

Box 1.
| Clinician targeted behaviors | Patient targeted behaviors |
| --- | --- |
| Use longitudinal PROM data in visit preparation and encounters. | Complete brief between-visit reflection on symptoms and progress. |
| Adjust treatment intensity proactively, including dose, visit frequency, and supportive services. | Raise concerns earlier when risk increases. |
| Use monitoring data as a collaborative tool rather than a control mechanism. | Participate more confidently in medication and care-intensity discussions. |

#### 1. Structured Patient-Reported Outcome Monitoring

Guided by a systematic review of single-item PROMs for substance use treatment, we prioritized constructs such as craving, treatment readiness, and self-efficacy, which demonstrated the strongest psychometric support and feasibility for routine monitoring in MBC.^24^ Patients complete brief PROMs at regular intervals between clinic visits, typically weekly during early treatment and then every 2–4 weeks once they have been more stable in care. Measures were selected to capture modifiable, clinically actionable aspects of bup-nx treatment and recurrent use risk: withdrawal symptoms, craving, medication adherence, medication side effects, opioid overdose risk, other substance use, global treatment improvement, and recovery community connection. We used single-item questions to minimize response burden while enabling longitudinal tracking. The full item set, response options, and recall periods are provided in **Supplement Table 1**. In the current specification, PROMs are timed several days before scheduled visits during the initial personalization phase, with less frequent pre visit monitoring as patients transition into longer term, more stable care. PROM delivery is automated through the EHR and sent via secure text message or patient portal. Delivery mechanics reflect patient preferences expressed in the needs assessment: assessments are designed for smartphone completion in a few minutes, arrive four days before scheduled visits during the personalization phase and one week before visits during longer-term monitoring so responses can inform shared visit preparation, and are followed by a reminder if not completed. For each aspect (for example, craving), individual responses are mapped to qualitatively defined severity bands (for example, normal, abnormal, critical) based on a combination of existing evidence, clinical judgment, and user feedback, and these bands inform downstream feedback, outreach options, and clinical prompts. Cut-points were drafted from published data and validated scales where available and from clinical guidance, then reviewed by the clinical members of the study team and adjusted during co-design when clinicians judged bands over- or under-inclusive. Because empirically validated thresholds do not exist for most single-item measures in this population, the bands are intentionally provisional and configurable. Responses near band boundaries default to the lower-intensity pathway, flagging for discussion at the next visit rather than generating outreach options. Clinician-facing materials frame banded scores as prompts to be interpreted against trends and the full clinical picture rather than as fixed cut-offs. The distribution of responses across bands, the frequency of outreach offers, and clinician overrides will be monitored during feasibility testing so that bands can be recalibrated. Higher severity bands increase the likelihood that responses will generate between visit outreach options or more intensive clinical consideration, but they do not by themselves mandate specific treatment changes; final decisions are made collaboratively by clinicians and patients, with prompts serving as non directive guidance.

#### 2. Automated Patient Feedback and Self-Reflection

Immediately after completing PROMs, patients receive brief, tailored feedback based on their responses. Feedback is designed to normalize common experiences, reinforce protective behaviors, and encourage reflection on symptoms or concerns that may warrant discussion. For patient-facing feedback, we drafted brief, non-punitive messages grounded in our prior qualitative work on trust-sensitive monitoring and in principles from motivational interviewing and recovery-oriented care (e.g., normalizing common experiences, reinforcing protective behaviors, and explicitly thanking patients for honesty). During the needs-assessment interviews, patients (n = 9) viewed example PROMs and feedback messages and provided brief verbal reactions. Their comments were used to refine wording to avoid punitive framing and to emphasize shared preparation for upcoming appointments. Draft messages were then reviewed alongside items during the clinician co-design sessions to ensure they aligned with harm-reduction and agency-supportive practice. Importantly, feedback does not trigger automatic care changes or punitive actions. When responses indicate elevated risk (e.g., high craving, worsening withdrawal, overdose risk), patients are offered the option to request contact from the care team before their next scheduled visit. This opt-in mechanism is intended to preserve agency, reduce perceptions of surveillance, and align with qualitative findings about trust-sensitive monitoring.

#### 3. Clinician-Facing Summaries and Decision Support

Patient responses are summarized in the EHR in formats designed for efficient clinical use, including simple visual trends over time and pre-populated visit note templates. **Figure 2** shows the EHR-embedded summary used to support longitudinal interpretation and collaborative decision-making. Clinicians can review longitudinal PROM data alongside medication history and toxicology results during visit preparation and in the encounter. Decision support is provided as non-directive prompts linking specific symptom patterns to potential clinical considerations, such as bup-nx dose adjustment, split dosing, formulation changes, changes in visit frequency, or additional supportive interventions. These prompts are framed as “guardrails” rather than algorithms; clinicians retain full discretion over care decisions. Underlying these prompts is an internal question-to-action map that links response bands within each domain to potential next steps, providing structured guidance while preserving clinician judgment and patient preferences. We predefined a set of candidate decision rules in the question-to-action map (e.g., severity thresholds for “normal/abnormal/critical,” conditions for branching to free text, and criteria for generating outreach prompts) and asked clinicians in each session to identify elements that felt unworkable, unclear, or misaligned with trust-preserving care. Proposed modifications were recorded in the map and carried forward to the next iteration.

**Figure 2.**
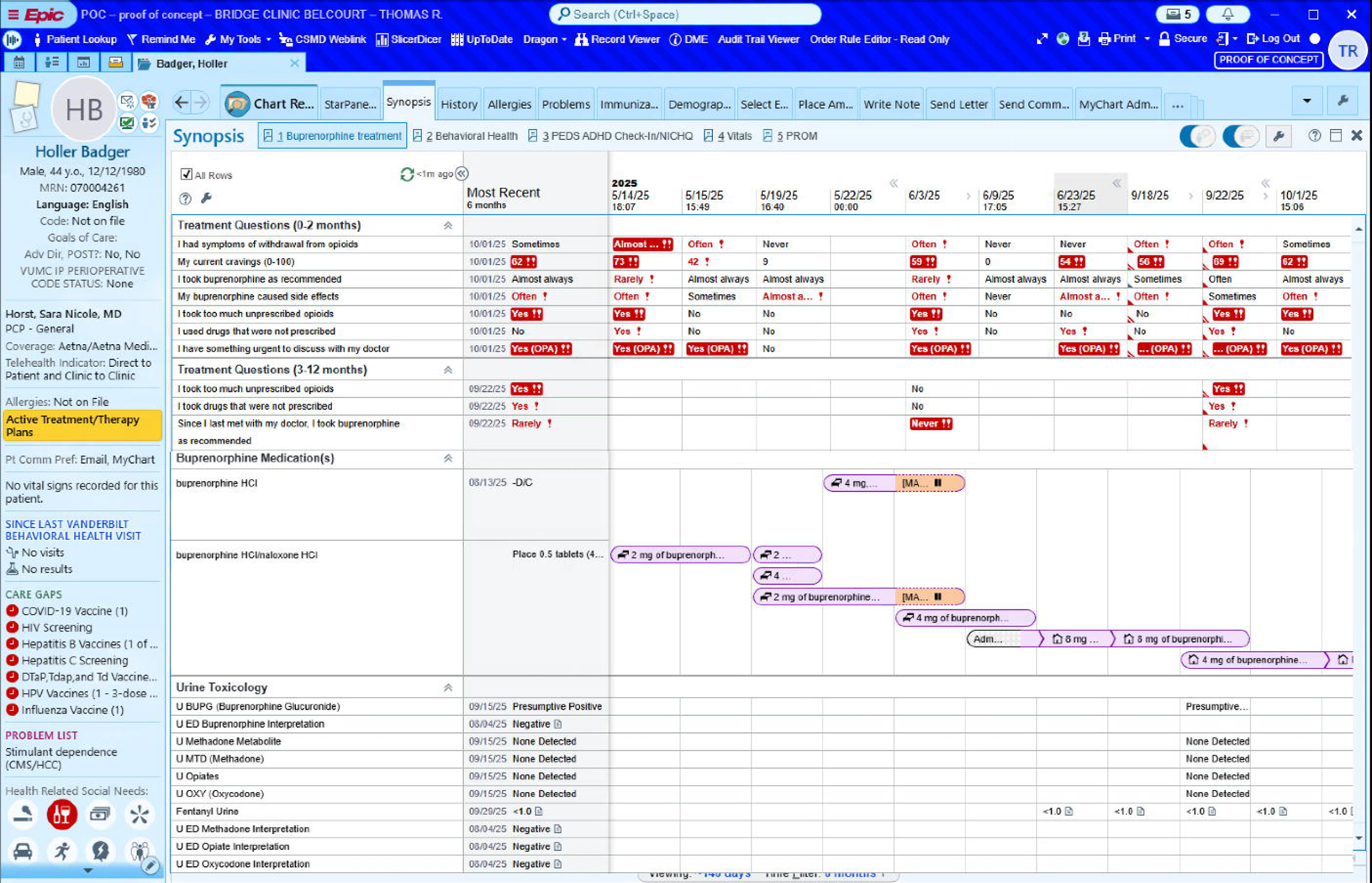
Clinician summary view for longitudinal assessment and decision making. (A) the longitudinal PROM trend rows with severity-banded responses, (B) the buprenorphine medication timeline, and (C) concurrent urine toxicology results

#### 4. Opt-In Between-Visit Outreach Pathway

Between-visit outreach occurs through existing clinic workflows and is triggered when patients explicitly request contact in response to feedback. Elevated responses do not automatically mandate outreach, reflecting a deliberate balance between safety monitoring and respect for patient readiness and agency. Outreach is conducted by members of the existing care team (e.g., nurses, social workers, or recovery coaches).

The exception to opt-in routing is a small set of prespecified critical responses, most importantly possible overdose. Overdose risk is screened with a single item asking, “since I last met with my doctor, I took too much unprescribed opioids or what I thought were opioids”, followed by a text box to complete the statement “when I took too much opioids, I…” An affirmative response is categorized as critical regardless of other answers. A critical response (1) displays patient-facing safety information, including overdose warning signs, naloxone guidance, and explicit instruction to call 911 in an emergency, together with a statement that responses are not monitored in real time; (2) asks the patient whether the concern can wait until the next visit; and (3) routes an EHR message to a designated clinic pool for review within 48 hours. Unlike other domains, critical responses generate team review even when the patient does not request outreach. This exception is explained to patients at enrollment so that mandatory safety follow-up is transparent rather than experienced as covert surveillance. A single item screen cannot capture the full complexity of overdose risk; it is intended as minimally burdensome trigger for human follow-up.

### Integration into clinical workflow and fidelity considerations

The intervention was designed to integrate into existing outpatient workflows without requiring additional in-person visits. PROM assignment, electronic delivery, scoring, feedback generation, and EHR summary creation are automated to minimize manual work. **Figure 3** shows the workflow for PROM completion and integration in decision making. Clinicians are expected to review PROM summaries primarily in conjunction with scheduled encounters or patient-initiated contact rather than continuously monitoring incoming data.

**Figure 3.**
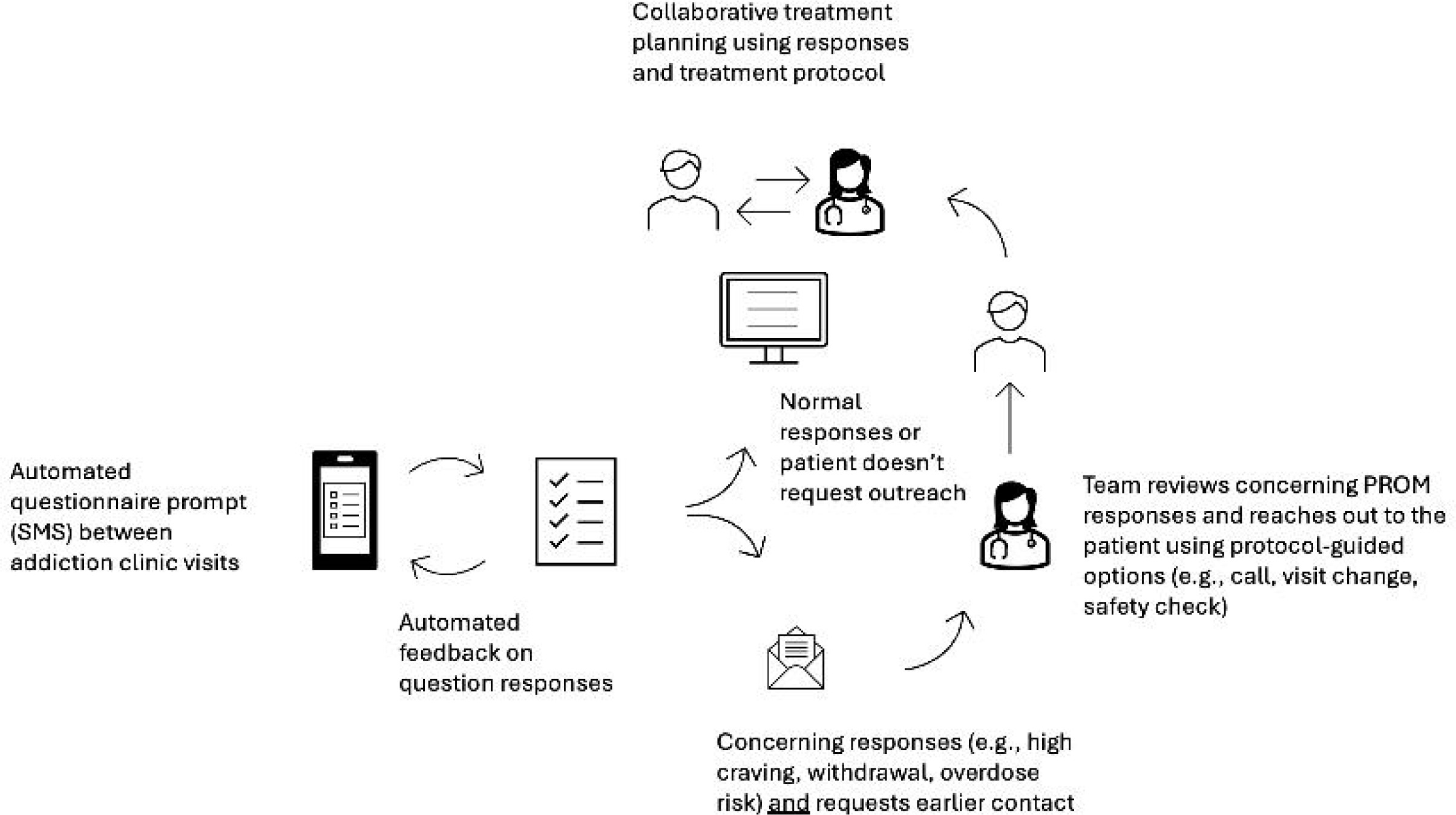
**Workflow for PROM completion and integration in treatment decisions, including the pathway for concerning responses that trigger clinician review and potential outreach.**

Planned fidelity indicators focus on: (1) patient engagement with PROMs (completion rates and patterns), (2) clinician access of PROM summaries within the EHR, and (3) reference to PROM data in visit documentation or discussions (as reflected in pre-populated note templates or chart review). These indicators define a minimum level of Bup MBC use, in which PROMs inform at least some aspect of conversation or decision making, and they will serve as core implementation outcomes in subsequent feasibility studies.

## RESULTS

Findings from the prior needs assessment and explicit user recommendations were translated into specific features of Bup MBC and implemented in the current intervention configuration. User input led to several pivotal refinements: recall periods were shifted from a fixed 7 day frame to a since last visit frame to better match clinical decision making; response scaling and severity banding, particularly for craving, were broadened to allow more nuanced interpretation; clinician facing interpretation and treatment considerations were added to turn raw scores into concrete, nondirective guardrails; and routing logic was revised so that most abnormal responses now offer a patient initiated outreach option rather than triggering automatic messages, with direct messaging reserved for situations where patients request contact. **Table 1** summarizes the core needs and corresponding design responses, and the following sections describe how these refinements addressed concerns about burden, clarity, trust, and workflow fit.

**Table 1.** Empirically Derived Needs and Corresponding Measurement. -**Based Care Design Responses**

|  |  |
| --- | --- |
| **<u>Need</u>** | **<u>Design response</u>** |
| Dynamic, between-visit recurrent use risk → high-frequency brief PROMs early in treatment | Interviews highlighted drop-off risk during transitions in visit spacing (for example, biweekly to monthly) and evolving risk between encounters; in response, we specified weekly PROMs during early “personalization” phase, followed by less frequent assessments later in care. |
| Fluctuating craving and under-detection → prioritized single-item craving measure with severity bands and thresholds | Clinicians described craving (even without withdrawal) as a key driver of recurrent use risk and dosing decisions; in response, we prioritized a craving slider with severity bands and escalation logic. |
| Withdrawal and pain narrow perceived options for coping and staying in treatment → withdrawal item + dose adequacy prompts | Patients described withdrawal as intolerable and as narrowing their perceived options for avoiding recurrent use and staying engaged in care; in response, the withdrawal assessment is tied to structured clinical considerations (dose adequacy, split dosing, formulation changes, and brief education about sublingual administration and factors that affect absorption). |
| Dose uncertainty and hesitancy to request changes → symptom-to-discussion prompts (“guardrails”) | Patients expressed uncertainty about dose adequacy, and clinicians noted underdosing risk; in response, clinician-facing prompts link symptom profiles (withdrawal, craving, adherence, side effects) to discussion topics and possible adjustments, functioning as nondirective guardrails rather than prescribing rules. |
| Fear of punitive monitoring and trust rupture → non-punitive feedback language | Monitoring could strengthen or rupture trust depending on framing; in response, patient-facing feedback emphasizes normalization, safety, and collaboration, and does not trigger automatic penalties. |
| Readiness fluctuates; motivation cannot be imposed → opt-in outreach by default | Both patients and clinicians emphasized agency, readiness, and harm reduction; in response, outreach is primarily patient-initiated when concerns cannot wait for the next visit (opt-in), rather than automatic escalation for all abnormal responses. |
| Safety-critical boundaries → mandatory escalation only for critical items | Safety concerns (for example, possible overdose) require timely clinician action; in response, overdose and other prespecified critical signals trigger urgent workflows and clinical follow-up guidance. |
| Visit time is limited; clinicians want workflow fit → EHR-embedded summaries + note template | Clinicians reported needing usable information “in the workflow”; in response, we implemented a longitudinal summary view and a visit note template with pre-populated responses and suggested considerations. |
| High burden from free-text reporting → branching only when needed | To limit burden while still capturing context, text boxes appear only after abnormal responses (for example, “describe withdrawal symptoms”). |

Note: Arrows indicate “if-then” relationships, where each empirically derived need led directly to the corresponding design response.

**Table 2.**
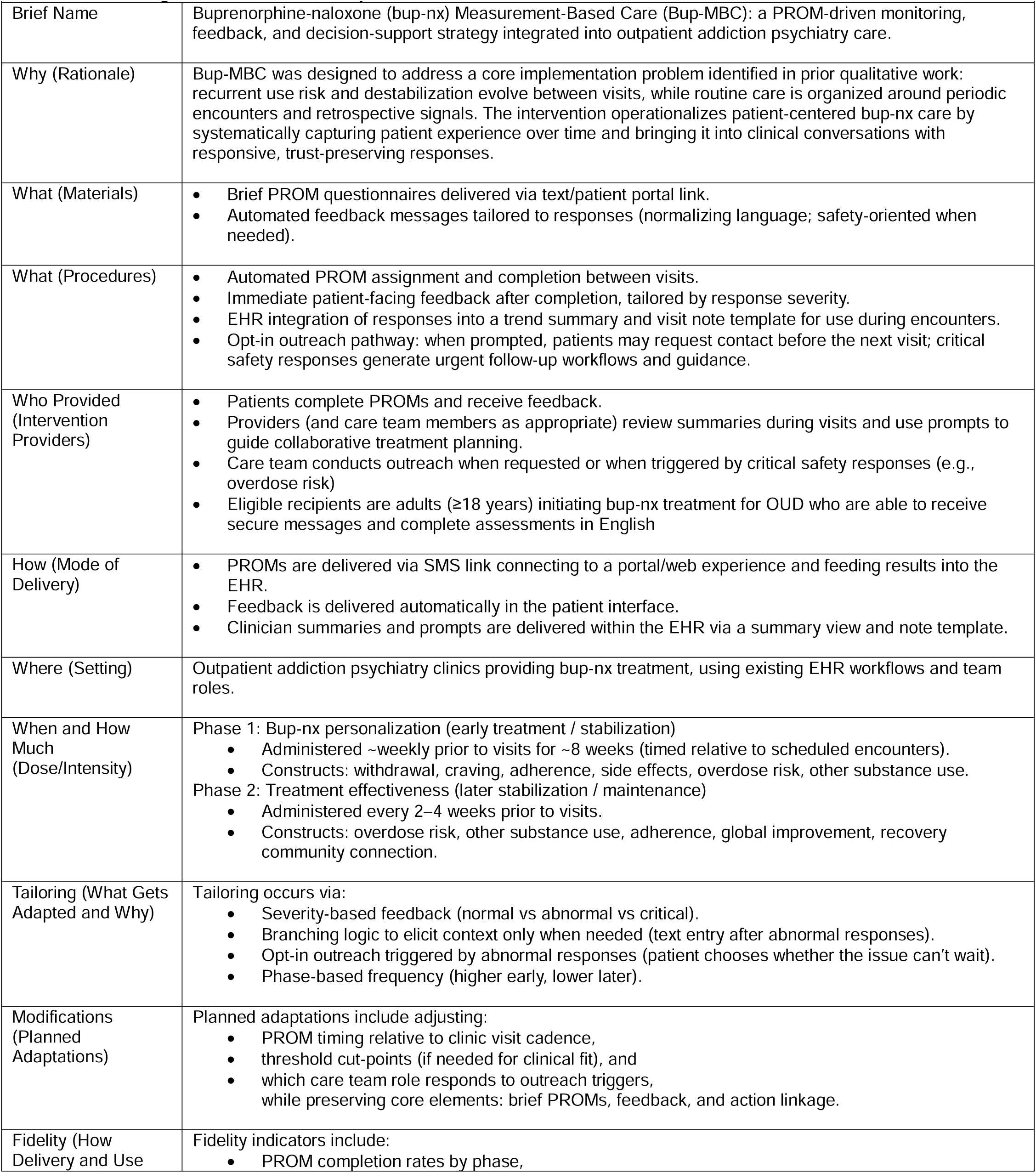
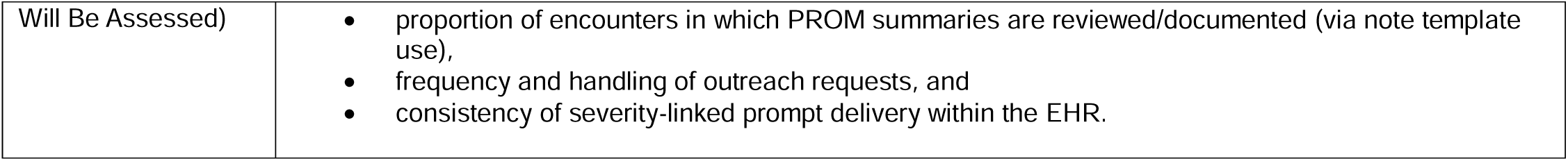
TIDieR-aligned intervention description.

### Refinements across co-design cycles

Across the three cycles, clinicians proposed 42 discrete changes; 27 were implemented as suggested, 10 were implemented with modifications, and 5 were not adopted (e.g., conflict with technical constraints of the EHR build). Cycle 1 concentrated on wording and recall periods, including the shift from a fixed 7-day recall to since-last-visit frame; Cycle 2 on response scaling, severity bands, and feedback language, including broadening the craving scale severity banding and removing or softening language patients had flagged; and Cycle 3 on clinician-facing interpretation, routing logic, and workflow fit, including converting most automatic outreach into a patient-initiated option and adding treatment considerations to the EHR summary and note template (**Supplement Table 3**).

### Addressing fluctuating recurrent use risk

Patients and clinicians described recurrent use vulnerability as continuous and evolving, with particularly high risk early in treatment and around transitions in visit frequency. Providers identified “loss points” when visits moved from every two weeks to every four weeks, noting that *“the most frequent time period… I’ve seen patients have returned to use or just drop off… is going from that two-week appointment mark to that four-week appointment…”* (Provider 1). In response, the intervention applies a two-phase PROM schedule: weekly pre visit assessments during an initial personalization phase, followed by pre visit PROMs every 2–4 weeks during longer term care. This schedule operationalizes between-visit vulnerability while limiting long-term burden.

### Minimizing burden, preserving actionability

Participants emphasized that during periods of instability they had limited bandwidth for lengthy assessments and that what mattered was capturing what they were “facing” between visits. One patient noted, *“What I’m facing… it’s going to be real… more than [what we talked about last time]”* (Patient 3). To support repeated use, each domain (e.g., craving, withdrawal, side effects) is measured with a single item, with optional free-text prompts only when responses indicate concern. This structure keeps assessments brief while still permitting contextual detail when clinically relevant. Patients described feedback as “reassuring” when it acknowledged ongoing struggle and thanked them for honesty, and they highlighted phrases that felt judgmental, which were subsequently removed or softened.

### Craving as an early signal

Providers and patients described craving as a key driver of recurrent use risk and dose decisions, often persisting despite adequate control of withdrawal. As one provider reported, *“…how much buprenorphine I would prescribe is [whether] the patient still [has] craving…,”* and a patient observed, *“Even when you’re on Suboxone… you get cravings still”* (Provider 4, Patient 2). The intervention therefore includes a single item craving scale with graded severity bands (from minimal to very high) that support longitudinal tracking and define when a patient is offered outreach and when clinicians see higher severity prompts. Lower levels prompt monitoring and discussion at the next visit; higher levels trigger the option for patient-initiated outreach and clinician prompts to consider dose adjustment or additional support.

### Clarifying dose adequacy

Patients expressed uncertainty about dose sufficiency and hesitancy to request changes, while clinicians described a tendency toward underdosing and the need to interpret complex symptom patterns. One patient said, *“I don’t know if I’m at the maximum… I’ve never asked…,”* and a provider noted *“…a lot of people do tend to get underdosed…”* (Patient 2, Provider 2). To address this, clinician-facing summaries include structured interpretation and considerations for each PROM domain (withdrawal, craving, adherence, side effects, overdose risk, other substance use), keyed to response categories. Frequent withdrawal, for example, prompts reassessment of dose adequacy, adherence timing, and sublingual administration technique (including potential effects of smoking or acidic beverages on absorption), whereas frequent side effects prompt clarification of timing and consideration of formulation or dosing changes. These prompts function as guardrails for discussion, not automated prescribing rules.

### Mitigating fear of punishment

Patients described worries that honest disclosure could jeopardize bup-nx access, and clinicians recalled instances where monitoring felt punishing. One provider recounted that a patient *“felt very like I had caught him doing something bad and that I was punishing him…”* (Provider 1). In response, patient-facing feedback was designed to normalize common experiences, thank patients for honesty, and emphasize that information is used to prepare for collaborative decisions rather than to punish. Feedback appears immediately after PROM completion and remains supportive even when responses signal elevated risk.

### Balancing readiness and safety

Participants emphasized that engagement depends on internal readiness and that change *“can’t be forced.”* Providers described a harm-reduction posture that avoids coercive pursuit when motivation is low. The intervention therefore uses an opt-in outreach pathway: abnormal responses present an option for patients to request earlier contact, and selecting this option generates a message to the care team with PROM context. At the same time, prespecified high risk responses (e.g., possible overdose) trigger an outreach prompt that enables rapid team contact when the patient indicates the issue cannot wait for the next visit.

### Between-visit connection and longitudinal use of data

Clinicians highlighted missed appointments, urgent refill requests, and changes in scheduling as difficult-to-interpret moments, and patients stressed that changes between encounters are central to safe decision-making. To provide more structured between-visit signals, PROMs are delivered electronically between visits so patients can report symptoms and barriers ahead of appointments rather than relying solely on ad hoc urgent messages. PROM data populate a longitudinal summary view and a visit note template that pre-populates recent responses for efficient review during encounters. In longer term monitoring, the construct set was expanded to include recovery community connection to capture protective supports relevant to sustained stability. This design supports consistent, longitudinal use of patient-generated data in visits without requiring additional in-person appointments or continuous clinician monitoring. Operationally, this is implemented by making the outreach item conditional, appearing only after responses that meet predefined trigger criteria, thereby limiting unnecessary prompts while preserving rapid contact when elevated risk is disclosed.

## DISCUSSION

This development study specifies an EHR-integrated MBC implementation strategy that was deliberately designed to address empirically identified capability, opportunity, and motivation gaps in outpatient bup nx treatment. In Bup MBC, assessment timing is aligned with periods of heightened vulnerability, brief PROMs and branching text are used to capture clinically actionable signals with minimal burden, non punitive feedback and an opt in outreach pathway are intended to protect agency while still allowing safety overrides, and longitudinal PROM data are linked to symptom specific prompts and trend summaries that support collaborative decision making. Effects on trust, agency, burden, and responsiveness should be interpreted as hypothesized and whether the strategy achieves them is an empirical question for the planned feasibility work.

Our prior needs assessment showed that recurrent use vulnerability often shifts between scheduled visits, that patients’ ability to exercise agency is bounded by physiologic and emotional constraints, and that monitoring can either strengthen or undermine trust depending on how information is used.^7^ The present strategy responds by making between visit fluctuations visible through brief PROMs, providing immediate non punitive feedback to patients, summarizing trends for clinicians in the EHR, and offering an outreach pathway that is largely opt in with clearly defined safety triggers. Taken together, these features position MBC not as a stand alone clinical intervention or automated decision system, but as a relational and behavioral scaffold that is intended to support safer, more collaborative bup nx treatment in routine care.

Our integrated development process, combining a systematic psychometric review, a theory informed qualitative needs assessment, and iterative co design, aligns with recommendations from prior PROM and implementation research in substance use treatment.^7,24–26^ This integrated development process is consistent with human centered design principles, in which interventions are iteratively shaped by end users’ needs, constraints, and feedback within their real clinical context.^34,35^ We note, however, that direct participation in the iterative co-design cycles was limited to clinicians; patients shaped item and feedback content earlier, through the needs assessment, rather than through the co-design cycles themselves. Patient perspectives entered the design indirectly through the needs assessment and reactions to draft materials. We also explicitly mapped empirically identified needs to concrete design features, consistent with calls for theory and data informed intervention development.^36^ Each design feature maps to a specific empirically identified need, as detailed in the Results and Table 1—for example, a two-phase PROM schedule for between-visit “loss points,” a graded craving measure, symptom-linked dosing prompts, and non-punitive, opt-in routing with narrow safety-critical triggers.^37,38^ This transparent mapping illustrates a theory- and data-informed approach to intervention development.

Opt-in outreach is intended to respect readiness and reduce perceptions of surveillance, but some patients with elevated symptoms may choose not to request contact; predefined safety signals therefore trigger a rapid outreach prompt (and site-specific escalation workflows when needed) for acute concerns. The choice of single-item measures emphasizes feasibility and longitudinal use but requires clear justification relative to multi-item validated scales and careful specification of how responses will guide care. The intervention assumes that clinicians will routinely review and use summaries in visits, which must be tested in practice.

### Limitations

First, although the overall process drew on human-centered design methods, patients did not participate directly in the iterative co-design cycles, with input limited to reactions on draft items and feedback messages during needs-assessment interviews. Second, current configuration assumes access to an SMS-capable phone or active patient portal account, adequate digital and health literacy, and English proficiency, which may concentrate benefit among already-advantaged patients as seen with other digital health tools.^39^ Third, agency is structurally constrained with the opt-in design assuming patients can and will self-advocate when risk rises. Fourth, the strategy assumes clinicians will routinely open and use the EHR summaries; non-use and alert burden are documented failure modes of clinical decision support.^40,41^ Fifth, the bands could be misinterpreted or misaligned, such as a “normal” response could be falsely reassuring to patients and clinicians, while a disclosed high-risk state might not be converted to contact under opt-in routing outside the prespecified critical triggers. Moreover, monitoring could still be experienced as surveillance despite non-punitive framing. The planned feasibility studies are designed to detect these failure modes, including null effects and harms, by assessing indicators such as reach and completion across patient subgroups, the distribution of responses, conversion of high-risk disclosures into contact, clinician summary access and documentation rates, and patient-reported experiences of the monitoring. Finally, this work is limited by its single academic setting, specific EHR infrastructure, and focus on one bup-nx treatment model, so although the underlying design principles may be transferable, exact workflows may not. The mechanisms proposed, including improved shared understanding, earlier identification of vulnerability, and more balanced responses, remain hypothesized.

At this pre-trial stage, key outcomes of interest are feasibility, acceptability, and preliminary impact on engagement, communication, and decision-making processes in routine bup-nx care. Subsequent feasibility and pilot studies will be needed to examine patient and clinician uptake, perceived usefulness, workflow fit, and potential effects on retention and crisis-driven care.

## CONCLUSION

This intervention development study specifies an EHR integrated measurement based care strategy designed to address empirically identified gaps in patient centered bup nx treatment. By aligning what is measured, when it is measured, and how it is used with patients’ and clinicians’ lived experience of recurrent use risk, bounded agency, and trust contingent monitoring, we designed a pragmatic, testable strategy that is ready for feasibility and pilot evaluation with the aim of making bup nx treatment safer and more collaborative in routine care.

## Data Availability

All data produced in the present study are available upon reasonable request to the authors and subject to data sharing agreements by the institution and participants.

## ACKNOWLEDGEMENTS

Primary funding for this study was provided by the Agency for Healthcare Research and Quality (AHRQ) through grant K08HS029695 (TJR).

## Notes

### Competing Interest Statement

The authors have declared no competing interest.

### Author Declarations

Vanderbilt University Medical Center Institutional Review Board gave ethical approval for this work.

## REFERENCES

1. Ferri M, Finlayson AJR, Wang L, Martin PR. Predictive factors for relapse in patients on buprenorphine maintenance. American Journal on Addictions. 2014;23(1):62–67. doi:10.1111/j.1521-0391.2013.12074.x

2. Marcovitz DE, McHugh RK, Volpe J, Votaw V, Connery HS. Predictors of early dropout in outpatient buprenorphine/naloxone treatment. American Journal on Addictions. 2016;25(6):472–477. doi:10.1111/ajad.12414

3. Singh V V., Dhawan A, Sarkar S, Mishra AK, Chadda RK. Relapse during opioid use disorder treatment: A pilot study to understand the reasons for opioid use during treatment. Ind Psychiatry J. 2023;32(2):361–368. doi:10.4103/ipj.ipj_87_22

4. Kelley AT, Incze MA, Baylis JD, et al. Patient-centered quality measurement for opioid use disorder: Development of a taxonomy to address gaps in research and practice. Subst Abus. Routledge. 2022;43(1):1286–1299. doi:10.1080/08897077.2022.2095082

5. Hichborn EG, Murray OB, Murphy EI, et al. Patient centered medication treatment for opioid use disorder in rural Vermont: a qualitative study. Addiction Science and Clinical Practice . 2025;20(1). doi:10.1186/s13722-024-00529-8

6. Cole ES, Allen L, Austin A, et al. Outpatient follow-up and use of medications for opioid use disorder after residential treatment among Medicaid enrollees in 10 states. Drug Alcohol Depend. 2022;241. doi:10.1016/j.drugalcdep.2022.109670

7. Reese T, Shah M, Wright A, et al. Balancing Return to Use Risk and Agency in Buprenorphine-naloxone Treatment: A Qualitative Needs Assessment to Inform Patient-Centered Care. medRxiv. Published online August 23, 2026. doi:10.64898/2026.08.21.26360804

8. Poulsen MN, Asdell PB, Berrettini W, McBryan K, Rahm AK. Application of the COM-B model to patient barriers and facilitators of retention in medication treatment for opioid use disorder in rural Northeastern United States: A qualitative study. SSM - Mental Health. 2022;2. doi:10.1016/j.ssmmh.2022.100151

9. Lewis CC, Boyd M, Puspitasari A, et al. Implementing Measurement-Based Care in Behavioral Health: A Review. JAMA Psychiatry. 2019;76(3):324–335. doi:10.1001/jamapsychiatry.2018.3329

10. He Y, Wang X, Wang Z, et al. Comparison of the Efficacy Between Standard Measurement-Base Care (MBC) and Enhanced MBC for Major Depressive Disorder: A Pilot Study. Neuropsychiatr Dis Treat. 2024;20:1465–1473. doi:10.2147/NDT.S468332

11. Forand NR, Nettiksimmons J, Brownell A, et al. The impact of measurement based care at scale: examining the effects of implementation on patient outcomes and provider behaviors. Frontiers in Health Services. 2025;5. doi:10.3389/frhs.2025.1659238

12. Marsden J, Tai B, Ali R, Hu L, Rush AJ, Volkow N. Measurement-based care using DSM-5 for opioid use disorder: can we make opioid medication treatment more effective? Addiction. 2019;114(8):1346–1353. doi:10.1111/add.14546

13. Ghitza UE. Research agenda evaluating measurement-based care for opioid use disorder among patients with co-occurring depressive disorders. Front Psychiatry. Frontiers Media SA. 2025;16. doi:10.3389/fpsyt.2025.1624642

14. Fortney JC, Unützer J, Wrenn G, et al. A tipping point for measurement-based care. American Psychiatric Association. 2017;68(2):179–188. doi:10.1176/appi.ps.201500439

15. Jamshidi N, Athavale A, Tremonti C, et al. Evaluation of adherence monitoring in buprenorphine treatment: A pilot study using timed drug assays to determine accuracy of testing. Br J Clin Pharmacol. 2023;89(7):1938–1947. doi:10.1111/bcp.15318

16. Farmer CM, Lindsay D, Williams J, et al. Practice guidance for buprenorphine for the treatment of opioid use disorders: Results of an expert panel process. Subst Abus. 2015;36(2):209–216. doi:10.1080/08897077.2015.1012613

17. Center for Substance Abuse Treatment. Clinical Guide Clinical Guidelines for the Use of Buprenorphine in the Treatment of Opioid Addiction. 2004. http://www.kap.samhsa.gov/products/

18. Atkins L, Francis J, Islam R, et al. A guide to using the Theoretical Domains Framework of behaviour change to investigate implementation problems. Implementation Science. 2017;12(1). doi:10.1186/s13012-017-0605-9

19. Huijg JM, Gebhardt WA, Crone MR, Dusseldorp E, Presseau J. Discriminant Content Validity of a Theoretical Domains Framework Questionnaire for Use in Implementation Research. 2014. http://www.implementationscience.com/content/9/1/11

20. Dyson J, Cowdell F. How is the Theoretical Domains Framework applied in designing interventions to support healthcare practitioner behaviour change? A systematic review. International Journal for Quality in Health Care. Oxford University Press. 2021;33(3). doi:10.1093/intqhc/mzab106

21. Oluwoye O, Fraser E. Barriers and Facilitators That Influence Providers’ Ability to Educate, Monitor, and Treat Substance Use in First-Episode Psychosis Programs Using the Theoretical Domains Framework. Qual Health Res. 2021;31(6):1144–1154. doi:10.1177/1049732321993443

22. Cane J, O’Connor D, Michie S. Validation of the theoretical domains framework for use in behaviour change and implementation research. Implementation Science. 2012;7:37.

23. Michie S, van Stralen MM, West R. The behaviour change wheel: A new method for characterising and designing behaviour change interventions. Implementation Science. 2011;6(1). doi:10.1186/1748-5908-6-42

24. Reese TJ, Tindle HA, Bachmann J, et al. Patient-reported outcomes for monitoring substance use treatment: A systematic review of single-item measures. Addiction. Published online 2026. doi:10.1111/add.70424

25. Migchels C, Zerrouk A, Crunelle CL, et al. Patient Reported Outcome and Experience Measures (PROMs and PREMs) in substance use disorder treatment services: A scoping review. Drug Alcohol Depend. Elsevier Ireland Ltd. 2023;253. doi:10.1016/j.drugalcdep.2023.111017

26. Okrant E, Reif S, Horgan CM. Development of an addiction recovery patient-reported outcome measure: Response to Addiction Recovery (R2AR). Subst Abuse Treat Prev Policy. 2023;18(1). doi:10.1186/s13011-023-00560-z

27. Rush AJ, Gore-Langton RE, Bart G, et al. Tools to implement measurement-based care (MBC) in the treatment of opioid use disorder (OUD): toward a consensus. Addiction Science and Clinical Practice. 2024;19(1). doi:10.1186/s13722-024-00446-w

28. Karnik NS, Marsden J, McCluskey C, et al. The opioid use disorder core outcomes set (OUD–COS) for treatment research: findings from a Delphi consensus study. Addiction. 2022;117(9):2438–2447. doi:10.1111/add.15875

29. Hoffmann TC, Glasziou PP, Boutron I, et al. Better reporting of interventions: Template for intervention description and replication (TIDieR) checklist and guide. BMJ (Online). 2014;348. doi:10.1136/bmj.g1687

30. Staniszewska S, Brett J, Simera I, et al. GRIPP2 reporting checklists: Tools to improve reporting of patient and public involvement in research. Res Involv Engagem. 2017;3(1). doi:10.1186/s40900-017-0062-2

31. Marcovitz DE, White KD, Sullivan W, et al. Bridging Recovery Initiative Despite Gaps in Entry (BRIDGE): study protocol for a randomized controlled trial of a bridge clinic compared with usual care for patients with opioid use disorder. Trials. 2021;22(1):757. doi:10.1186/s13063-021-05698-4

32. Kast KA, Le TD V., Stewart LS, et al. Impact of inpatient addiction psychiatry consultation on opioid use disorder outcomes. Am J Addict. Published online March 28, 2024. doi:10.1111/ajad.13540

33. Reddy IA, Audet CM, Reese TJ, Peek G, Marcovitz D. Provider Perceptions toward Extended-Release Buprenorphine for Treatment of Opioid Use Disorder. J Addict Med. Published online September 3, 2024. doi:10.1097/adm.0000000000001320

34. Göttgens I, Oertelt-Prigione S. The Application of Human-Centered Design Approaches in Health Research and Innovation: A Narrative Review of Current Practices. JMIR Mhealth Uhealth. JMIR Publications Inc. 2021;9(12). doi:10.2196/28102

35. Lyon AR, Aung T, Bruzios KE, Munson S. Human-Centered Design to Enhance Implementation and Impact in Health. Public Health. 2026;44:467–485. doi:10.1146/annurev-publhealth

36. Teck JTW, Gittins R, Zlatkute G, Pérez AO, Galea-Singer S, Baldacchino A. Developing a Theoretically Informed Implementation Model for Telemedicine-Delivered Medication for Opioid Use Disorder: Qualitative Study With Key Informants. JMIR Ment Health. 2023;10(1). doi:10.2196/47186

37. Williams BE, Martin SA, Hoffman KA, Andrus MD, Dellabough-Gormley E, Buchheit BM. “It’s within your own power”: shared decision-making to support transitions to buprenorphine. Addiction Science and Clinical Practice . 2025;20(1). doi:10.1186/s13722-025-00555-0

38. Okrant E, Reif S, Bailey GL, et al. Using a quality improvement framework to evaluate the feasibility of implementing a patient-reported outcome measure for recovery in an office-based treatment setting for opioid use disorder. Addiction Science & Clinical Practice. Published online December 22, 2025. doi:10.1186/s13722-025-00632-4

39. Lyles CR, Wachter RM, Sarkar U. Focusing on Digital Health Equity. JAMA - Journal of the American Medical Association. American Medical Association. 2021;326(18):1795–1796. doi:10.1001/jama.2021.18459

40. Ancker JS, Edwards A, Nosal S, Hauser D, Mauer E, Kaushal R. Effects of workload, work complexity, and repeated alerts on alert fatigue in a clinical decision support system. BMC Med Inform Decis Mak. 2017;17(1):1–9. doi:10.1186/s12911-017-0430-8

41. Reese TJ, Kawamoto K, Fiol G Del, et al. When an Alert is Not an Alert : A Pilot Study to Characterize Behavior and Cognition Associated with Medication Alerts. AMIA Annual Symposium Proceedings. 2018;2018:1488–1497.

