## Supplement Table 1, Supplement Table 2, Supplement Table 3 for "A Measurement-Based Care Strategy for Buprenorphine-Naloxone Treatment (Bup-MBC): Development of an EHR-Integrated Intervention"

| **Supplement Table 1. Phase 1 – Bup-nx personalization.** | | | | | |
| --- | --- | --- | --- | --- | --- |
| **Response option** | **Severity** | **Patient-facing feedback** | **Branching prompt** | **Clinical interpretation** | **Clinician-facing treatment considerations** |
| **Withdrawal (item 1).** *“Since I last met with my doctor, I have had symptoms of withdrawal from opioids (restlessness, vomiting, diarrhea, or whole body aches etc.)”* | | | | | |
| Never | **Normal** | Your symptoms are well controlled. Keep up the good work! | — | None or minimal withdrawal symptoms | Maintain current buprenorphine dose and regimen. |
| Rarely | **Normal** | Mild symptoms can happen. Track any changes and share them at your next visit. | — | Mild withdrawal symptoms | Maintain current dose and regimen; reassure patient. |
| Sometimes | **Normal** | Noticing some symptoms – make a quick note and share with your care team for support at your next visit. | — | Noticeable withdrawal symptoms | Evaluate dose adequacy; consider small dose increase (e.g., +2 mg). Assess adherence and dose timing; consider split dosing if symptoms cluster at specific times. Reevaluate comorbidities or concomitant substance use affecting withdrawal. |
| Often | **Abnormal** | Frequent symptoms – please go back to the question and describe them (if you haven’t) and a member of your care team will contact you. | *“My withdrawal symptoms are…” (text box); triggers outreach item* | Frequent withdrawal symptoms | Increase dose in 2–4 mg increments as tolerated. Strongly assess adherence and missed doses; consider split dosing. Evaluate for drug interactions or other factors affecting withdrawal. |
| Almost always | **Critical** | Persistent symptoms – please go back to the question and describe them (if you haven’t) and a member of your care team will contact you. | *“My withdrawal symptoms are…” (text box); triggers outreach item* | Persistent withdrawal symptoms | Consider larger dose increase, up to maximum recommended (e.g., 24–32 mg daily). Assess need to switch to long-acting injectable formulation. Intensify clinical monitoring and support. |
| **Craving (item 2).** *“Place the circle on the line where your current cravings are (0–100 visual analog slider; “No craving at all” to “Strongest craving ever”)”* | | | | | |
| 0–10 | **Normal** | Your cravings are extremely well controlled. Keep up the good work! | — | None or minimal craving | Maintain dose. Reinforce adherence and sublingual technique (dissolve fully; no food or drink until dissolved). Monitor routinely. |
| 11–30 | **Normal** | Mild cravings – this is common. Keep tracking your cravings and share any changes at your next visit. | — | Mild craving | Assess triggers. Confirm sublingual technique/absorption before any change (dissolve fully; avoid food or drink, especially acidic, until dissolved). Small dose increase if craving persists. |
| 31–50 | **Abnormal** | You’re having some cravings now – take note and talk with your care team at your next visit about how you’re managing. | — | Moderate craving | Confirm adherence and sublingual absorption before escalating (poor technique can mimic underdosing). If optimal, increase 2–4 mg or split dosing; address precipitating factors. |
| 51–70 | **Critical** | High cravings – Your medication or support might need adjustment. A member of your care team will contact you. | *Triggers outreach item* | High craving | Confirm adherence and sublingual technique. If adequate, increase 2–4 mg up to recommended maximum. Add behavioral support. |
| 71–100 | **Critical** | Very strong cravings – Your medication or support might need adjustment. A member of your care team will contact you. | *Triggers outreach item* | Very high craving | Verify technique, then maximize dose per guidelines (e.g., 24–32 mg). Intensify support; consider extended-release injectable (bypasses sublingual absorption). |
| **Adherence (item 3).** *“Since I last met with my doctor, I took buprenorphine as recommended”* | | | | | |
| Never | **Critical** | It looks like you haven’t been able to take your medication. If you haven’t, please go back to the question and describe how we can help you and a member of your care team will contact you. | *“My buprenorphine is hard to take because…” (text box); triggers outreach item* | Complete buprenorphine nonadherence | Assess barriers immediately (access, cost, stigma, side effects). Intensify follow-up; consider daily supervised or directly observed dosing. Strongly consider transition to long-acting injectable buprenorphine (LAIB). Provide education and motivational interventions. |
| Rarely | **Abnormal** | It’s been tough to take your medication. If you haven’t, please go back to the question and describe how we can help you and a member of your care team will contact you. | *“My buprenorphine is hard to take because…” (text box); triggers outreach item* | Very poor buprenorphine adherence | Detailed assessment of barriers. Consider LAIB to remove daily burden. Enhance reminders/support (calls, texts); involve case management/social supports. |
| Sometimes | **Abnormal** | Taking your medicine can be challenging. If something is getting in your way, please take note and talk with your care team at your next visit. | *Triggers outreach item* | Partial/intermittent buprenorphine adherence | Clarify pattern of missed doses (times, triggers). Provide adherence tools (reminders, pill boxes); explore LAIB option. Schedule more frequent follow-up; address contributing factors (side effects, forgetfulness, motivation). |
| Often | **Normal** | Great job taking your medication most of the time! If anything is making it hard, please take note and talk with your care team at your next visit. | — | Good but not perfect buprenorphine adherence | Reinforce positives; problem-solve missed doses and clarify their impact. Consider behavioral supports or technology-based reminders; LAIB may be offered for convenience. |
| Almost always | **Normal** | Excellent work staying on track with your medication. Keep it up! | — | Near-complete buprenorphine adherence | Maintain current approach; reinforce adherence and discuss occasional lapses. Routinely monitor; discuss LAIB if patient prefers more convenience. |
| **Side effects (item 4).** *“Since I last met with my doctor, my buprenorphine caused side effects that bothered me (headache, nausea, constipation)”* | | | | | |
| Never | **Normal** | You’re doing well with your medication. Great job! | — | No bothersome side effects | Maintain current dose and regimen; continue standard monitoring and education. |
| Rarely | **Normal** | You are having a few mild side effects. Keep tracking them, and let us know if things change at your next visit. | — | Minimal/mild side effects | Maintain dose; ask about specific side effects. Provide reassurance and education; monitor at next visit. |
| Sometimes | **Normal** | You are having some side effects. Please keep track and talk with your care team if they get worse at your next visit. | — | Noticeable side effects | Inquire about type, timing, and severity. Consider minor dose reduction or split dosing; manage specific side effects with adjunctive medications or lifestyle modification. Assess drug–drug interactions and adherence. |
| Often | **Abnormal** | You’re having side effects more often than we expect. If you haven’t, please go back to the question and describe them and a member of your care team will contact you. | *“The side effects I have are…” (text box); triggers outreach item* | Moderate side effects | Detailed side-effect review. Reduce total daily dose if possible or split dosing; switch formulation (film, tablet, or LAIB) if linked to delivery method. Treat specific side effects directly; increase monitoring and follow-up frequency. |
| Almost always | **Abnormal** | You’re having side effects most of the time. If you haven’t, please go back to the question and describe them and a member of your care team will contact you. | *“The side effects I have are…” (text box); triggers outreach item* | Severe/persistent side effects | Consider significant dose reduction or prompt transition to alternative formulation (e.g., LAIB). Intensive clinical management; refer to specialty care if side effects are dangerous or intolerable. Urgently manage any serious adverse effect; provide safety education. |
| **Opioid use and overdose (item 5; safety-critical).** *“Since I last met with my doctor, I took too much unprescribed opioids or what I thought were opioids”* | | | | | |
| Yes | **Critical** | Thank you for being honest. For your safety, please go back to the question (if you haven’t) and describe what happened and a member of your care team will contact you to help. | *“When I took too much opioids, I…” (text box); triggers outreach item* | Possible opioid overdose | Assess immediate safety (timing, severity, need for medical care, naloxone use). Increase buprenorphine dose if indicated, up to recommended maximum (typically 24–32 mg daily). Intensify monitoring (clinical contact, urine drug screens, follow-up). Review and reinforce overdose prevention; offer naloxone kit. Address underlying causes (inadequate dose, stressors, mental health). Consider LAIB if adherence poor or risk high; refer to or coordinate with specialty addiction services for recurrent overdose. Schedule rapid return visits. |
| No | **Normal** | You’re staying on track – great job! If you have any problems or concerns, please let your care team know at your next visit. | — | No opioid use, or not perceived as a problem | Maintain current buprenorphine dose and regimen. |
| **Other substance use (item 6).** *“Since I last met with my doctor, I used unprescribed sedatives (benzos, downers, or alcohol), unprescribed stimulants (methamphetamine or cocaine), or other drugs that were not prescribed”* | | | | | |
| Yes | **Abnormal** | Thank you for sharing this. Using other drugs can increase risks. If you haven’t, please go back to the question and tell us more about your use and how we can support you – someone from your care team will follow up to help. | *“The other drug(s) I used were…” (text box); triggers outreach item* | Possible other drug use | Assess immediate safety risks (evaluate overdose risk with sedatives or alcohol). Review drug interactions (sedatives increase overdose risk with buprenorphine). Discuss behavioral risk factors, triggers, and context. Enhance monitoring (visits, urine drug screens, check-ins); offer referral for concurrent substance use treatment or behavioral support. Typically maintain buprenorphine dose to support OUD stabilization; consider LAIB if ongoing unsafe use despite optimization. Update safety and overdose prevention plans (naloxone, harm-reduction counseling); document findings. |
| No | **Normal** | Great job avoiding other drugs. Keep up the good work! If you have concerns or need support, let your care team know at your next visit. | — | No other drug use, or not perceived as a problem | Maintain current buprenorphine dose and regimen. |
| **Outreach (item 7; shown only when a triggering response occurs above).** *“I have something urgent to discuss with my doctor before my visit”* | | | | | |
| Yes | **Critical** | A member of your care team will contact you. | *“I need to discuss my…” (text box, required)* | Urgent item to discuss | Secure in-basket message routed to the care team for between-visit outreach. |
| No | **Normal** | — | — | Abnormal responses can wait until the next appointment | Responses remain visible in the EHR summary for review at the next visit. |

Delivered 4 days before each scheduled addiction psychiatry visit for approximately 8 weeks (typically weekly)

| **Supplement Table 2. Phase 2 – Treatment effectiveness.** | | | | | |
| --- | --- | --- | --- | --- | --- |
| **Response option** | **Severity** | **Patient-facing feedback** | **Branching prompt** | **Clinical interpretation** | **Clinician-facing treatment considerations** |
| **Global impression of improvement (item 4).** *“Compared to how I was before starting treatment, my drug problems are”* | | | | | |
| Very much better | **Normal** | Fantastic to see so much improvement! Keep doing what works for you. | — | Excellent improvement; remission or near-remission | Maintain current regimen; reinforce adherence. Consider spacing out visits if stable over a prolonged period. Continue regular monitoring, recurrence prevention, and patient empowerment. |
| Much better | **Normal** | You’re making strong progress – well done! Let’s keep building on this together. | — | Sustained substantial improvement | Maintain current approach with positive reinforcement. Review for residual symptoms; regular but possibly less frequent monitoring. Encourage ongoing engagement and recurrence prevention. |
| A little better | **Normal** | Glad you’re seeing improvement. If you have any concerns or want more support, let us know. | — | Meaningful but partial improvement | Evaluate for further optimization: reassess persistent withdrawal, craving, or side effects. Consider minor buprenorphine dose adjustment; enhance behavioral supports or address social determinants. |
| No change | **Normal** | Change can take time. Please share what’s working or not – we’re here to help at your next visit. | — | Lack of meaningful clinical response | Systematic review: assess adherence, dose adequacy, and barriers. Increase dose if withdrawal/craving present; intensify behavioral/psychosocial interventions. Screen for comorbidities and interactions; consider LAIB; monitor closely. |
| A little worse | **Abnormal** | Sometimes recovery is hard. Tell us what’s making things difficult so we can support you. | *“Things that have made my situation worse include…” (text box)* | Worsening problems despite treatment | Prompt intervention: comprehensive reassessment (adherence, dose, substance use, mental health). Escalate dose if indicated; address new triggers/stressors. Consider switch to LAIB; intensify behavioral and social supports; increase contact frequency. |
| Much worse | **Abnormal** | We want your treatment to work better. We should talk about making changes to help you feel better. | *“Things that have made my situation worse include…” (text box); triggers outreach item* | Substantial clinical deterioration | Urgent reassessment: immediate review for ongoing opioid use, overdose risk, and adherence. Maximize buprenorphine dose (up to 24–32 mg); consider LAIB or change in setting (higher level of care). Rapidly mobilize multidisciplinary supports; ensure safety and harm reduction (naloxone, overdose prevention). |
| Very much worse | **Abnormal** | Your health and safety matter. We need to work together to find options that will help you right away. | *“Things that have made my situation worse include…” (text box); triggers outreach item* | Major clinical decline; high risk | Immediate action: evaluate for hospitalization, crisis intervention, or acute stabilization. Consider transfer to specialty addiction or inpatient care. Maximize pharmacotherapy; use LAIB if feasible. Address imminent safety, overdose, and suicidality; involve family/support system urgently. |
| **Recovery community connection (item 5).** *“In the past month, I have felt like I’m part of a recovery community”* | | | | | |
| Never | **Abnormal** | Feeling connected can help recovery. Let’s talk about how to build support together at your next visit. | — | No sense of recovery community connection | Assess isolation risk. Discuss importance of peer support; encourage or refer to mutual aid groups, peer recovery support, or group therapy. Increase care team touchpoints; monitor for loneliness, recurrence risk, or disengagement. |
| Rarely | **Abnormal** | You’re not alone – let’s discuss ways to find more support at your next visit. | — | Very limited connection | Explore barriers to participation (transportation, stigma, scheduling, comfort). Provide information on accessible resources, including virtual/online recovery communities; consider peer navigators or recovery coaches. |
| Sometimes | **Normal** | Some community connection is a good start. Let’s explore ways to strengthen it together at your next visit. | — | Partial or intermittent involvement | Reinforce existing community linkages; problem-solve challenges to regular participation. Highlight benefits of greater connection; offer more structured group therapy if desired. |
| Often | **Normal** | Great job joining and staying active in your recovery community! How has this helped you? | *“I have participated in…” (text box)* | Strong community involvement | Recognize and praise engagement; encourage continued involvement. Explore how involvement supports recovery; maintain current treatment plan. |
| Almost always | **Normal** | Wonderful – being part of a recovery community really helps. Keep building those connections! | *“I have participated in…” (text box)* | Full, consistent sense of connection | Reinforce the role of community connection; encourage leadership or sponsorship roles if appropriate. With consent, use the patient’s experience to mentor others; maintain regimen and continue supporting community-based recovery. |
| **Outreach (item 6; shown only when a triggering response occurs above).** *“I have something urgent to discuss with my doctor before my visit”* | | | | | |
| Yes | **Critical** | A member of your care team will contact you. | *“I need to discuss my…” (text box, required)* | Urgent item to discuss | Secure in-basket message routed to the care team for between-visit outreach. |
| No | **Normal** | — | — | Abnormal responses can wait until the next appointment | Responses remain visible in the EHR summary for review at the next visit. |

Delivered 1 week before addiction psychiatry visits for 12 months or until buprenorphine discontinuation (typically every 2–4 weeks). All items are required. Items 1–3 (opioid use and overdose, other substance use, and adherence) are identical in wording, response options, severity bands, feedback, branching, and treatment considerations to the corresponding Phase 1 items (Supplement Table 1).

| **Supplement Table 3. Changes proposed and their disposition across the three co-design cycles.** | | | | | | |
| --- | --- | --- | --- | --- | --- | --- |
| **Cycle** | **Primary focus** | **Proposed** | **Implemented** | **Implemented with modification** | **Not adopted (rationale)** | **Example changes** |
| 1 | Item wording and recall periods | 24 | 15 | 8 | 1 (limited space for potential bup-nx side effects) | Shift from fixed 7-day recall to a since-last-visit frame |
| 2 | Response scaling, severity bands, and feedback language | 11 | 9 | 1 | 1 (validated categories for global impression of improvement) | Broadened 0–100 craving scale with five bands; softened language patients had flagged as judgmental |
| 3 | Clinician-facing interpretation, routing logic, and workflow fit | 7 | 3 | 1 | 3 (limited capability to send questionnaire precisely between visits; limited visualization options for measures over time in EHR summary; limited options to link medication order to considerations) | Most automatic outreach converted to a patient-initiated option; treatment considerations added to the EHR summary and note template |
